# SmokeBERT: A BERT-based Model for Quantitative Smoking History Extraction from Clinical Narratives to Improve Lung Cancer Screening

**DOI:** 10.1101/2025.06.18.25329870

**Authors:** Yiming Xue, Yunzheng Zhu, Luoting Zhuang, YongKyung Oh, Ricky Taira, Denise R. Aberle, Ashley E. Prosper, William Hsu, Yannan Lin

**Affiliations:** Department of Statistics & Data Science, University of California, Los Angeles, CA, USA; Medical & Imaging Informatics, Department of Radiological Sciences, David Geffen School of Medicine at UCLA, Los Angeles, CA, USA

## Abstract

Tobacco use is a critical risk factor for diseases such as cancer and cardiovascular disorders. While electronic health records can capture categorical smoking statuses accurately, granular quantitative details, such as pack years and years since quitting, are often embedded in clinical narratives. This information is crucial for assessing disease risk and determining eligibility for lung cancer screening (LCS). Existing natural language processing (NLP) tools excelled at identifying smoking statuses but struggled with extracting detailed quantitative data. To address this, we developed SmokeBERT, a fine-tuned BERT-based model optimized for extracting detailed smoking histories. Evaluations against a state-of-the-art rule-based NLP model demonstrated its superior performance on F1 scores (0.97 vs. 0.88 on the hold-out test set) and identification of LCS-eligible patients (e.g., 98% vs. 60% for ≥20 pack years). Future work includes creating a multilingual, language-agnostic version of SmokeBERT by incorporating datasets in multiple languages, exploring ensemble methods, and testing on larger datasets.

## Introduction

Quantitative smoking history (e.g., pack years and years since quitting) is crucial for various clinical applications, particularly lung cancer screening (LCS). The 2021 United States Preventive Services Task Force (USPSTF) recommended annual low-dose computed tomography screening for lung cancer among individuals with at least a 20 pack-year smoking history who currently smoke or have quit within the past 15 years.^1^ Accurate smoking information is essential for determining eligibility for LCS. Despite over a decade of clinical LCS programs in the United States following the 2013 USPSTF recommendations, the adoption rate of LCS among eligible individuals remains strikingly low, with a national average of 16% in 2022.^2^ To facilitate LCS activities, the National Committee for Quality Assurance (NCQA) is developing Healthcare Effectiveness Data and Information (HEDIS®) measures for LCS and Tobacco Use Screening and Cessation.^3, 4^ To support the implementation of these quality measures, there is an urgent need for tools capable of extracting quantitative smoking history from electronic medical records (EHRs). Studies have shown that unstructured, free-text clinical narratives in EHRs often serve as a more reliable source of information compared to structured data fields.^5^ However, the extraction of this information necessitates the use of natural language processing (NLP) tools.^6^

Existing natural language processing (NLP) tools like cTAKES^7^ and CLAMP^8^ have proven effective in identifying broad smoking statuses such as “current smoker” or “former smoker”, but they face significant challenges when it comes to extracting detailed quantitative information, such as pack-years and years since quitting. The state-of-the-art (SOTA) model for extracting information from clinical narratives is a rule-based NLP model developed using clinical notes from a single institution.^9^ Although the rule-based NLP model can achieve good performance of both F1-score (0.94 to 0.96) and recall (0.94 to 0.96) on in-house test data, the model lacks generalizability on new datasets, where physicians may have different writing styles for documenting quantitative smoking history. Moreover, the current SOTA model captures some quantitative smoking variables (i.e., pack years, packs per day, years smoked, and years since quitting) but is limited in scope, missing important variables that are slightly different from the essential ones, (e.g., cigarettes per day and quit in the year). While new variables can be accommodated by adding additional rules, this approach is not scalable, as the number of rules needed can become unmanageable, resulting in increased development time and potential conflicts among the rules.

Bidirectional Encoder Representations from Transformers (BERT)-based models have become the baseline for many NLP tasks due to their focus on the bidirectional context within the text, making them particularly effective for domain-specific tasks like clinical named entity recognition (NER). Compared to transformers, Generative Pre-trained Transformers (GPTs), or other large language models, selected BERT models excel in extracting smoking-related information with precision and contextual understanding tailored to healthcare and biomedical fields. These models benefit from domain-specific pretraining, which has been shown to offer significant improvements over continual adaptation of general-purpose language models.^10^ Additionally, conducting an additional stage of domain-adaptive pretraining has demonstrated performance gains across both high-and low-resource settings.^11^ By leveraging models like ClinicalBERT and BioBERT, which have already undergone this domain-specific adaptatio , we can achieve improved representation learning. We aim to develop generalizable and scalable models to extract clinical narratives that specify patient quantitative smoking history, leveraging BERT-based models^12^ on a multi-institutional dataset. We hypothesize that BERT-based models outperform the SOTA model in the F1 score for both NER and clinical tasks.

## Methods

This study was approved by the Institutional Review Board at the University of California, Los Angeles (UCLA).

### Datasets

Our study was conducted on two datasets: the smoking history dataset and the MIMIC-III dataset. The smoking history dataset consists of 3,261 sentences describing smoking history from clinical reports. A portion of the smoking history dataset was derived from the i2b2 challenge^13^, which includes 502 de-identified discharge summaries sourced from the Partners HealthCare system^14^. The remaining data were extracted from UCLA ophthalmology notes and radiology reports using keyword matching based on lexical variants of ‘smoke’ (e.g., smoking, smoked, smoker), ‘tobacco’, ‘cigarette’, and “pack-year”. In addition, MIMIC-III contains de-identified health records from over 58,000 hospital admissions, including approximately 2 million clinical notes, demographics, lab results, and more, widely used for healthcare analytics such as mortality prediction^15^ and blood pressure estimation^16^. A subset of 3,191 sentences was collected using keyword matching with a similar set of keywords containing lexical variants of ‘smoke’, ‘tobacco’, ‘cigarette’, and ‘pack-year’. Sentences from the MIMIC-III subset were also used for the clinical tasks to identify individuals eligible for LCS. **Table 1** lists the number of entities in each dataset.

**Table 1.**
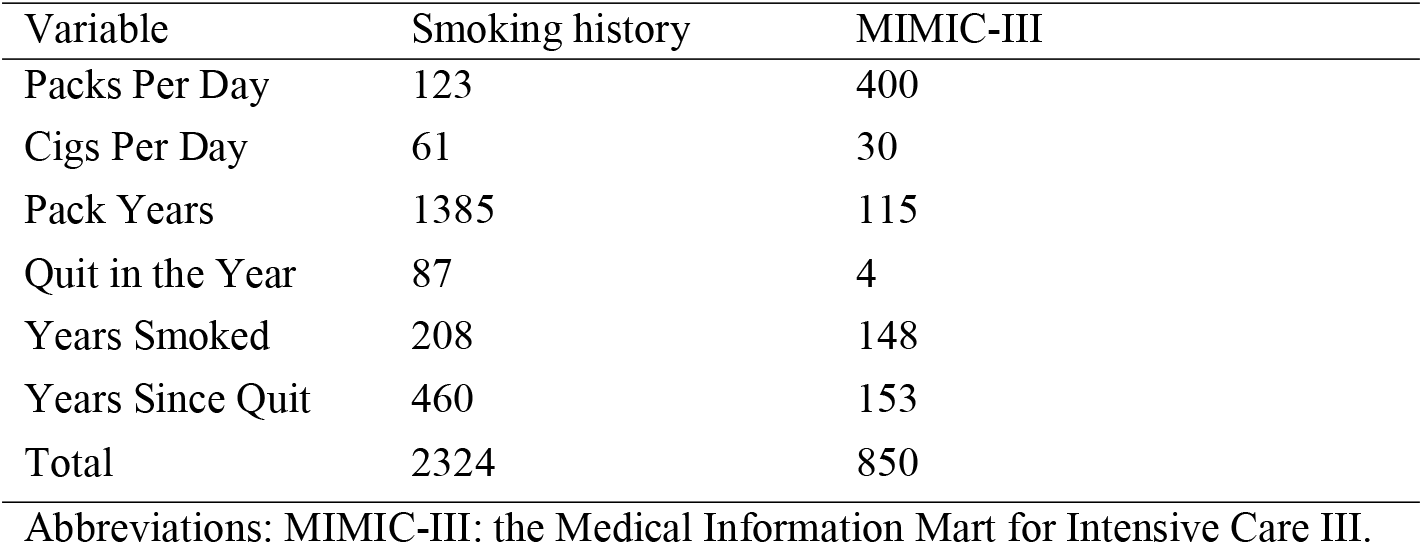
Number of entities in the smoking history and MIMIC-III Datasets.

### Data Annotations

Six variables were manually annotated on both datasets:”packs per day”,”cigarettes per day”,”years smoked”,”pack years”, “years since quitting”, and “quit in the year” using the NER Annotator for Spacy annotation tool.^17^**Figure 1** illustrates one example of the annotation. Two readers (Y.L. and Y.X.) annotated 300 randomly selected sentences from the smoking history dataset. Inter-rater agreement was described in the **Supplemental Methods**. Excellent reliability (two-way mixed effects model, single rater absolute intraclass correlation coefficient between 0.97 and 1) was achieved for all smoking variables (**Table S1** in the Supplement). Discrepancies were resolved before one of the annotators (Y.L.) completed the remaining annotations.

**Figure 1.**
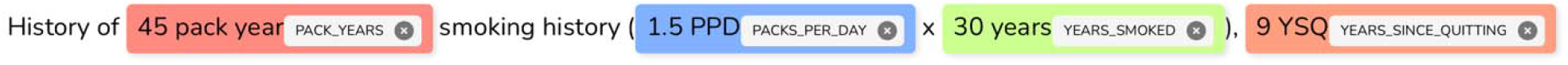
Manual annotation of smoking history from clinical narratives.

### Baseline Model

Our baseline model is a rule-based two-stage approach, which is the SOTA model for quantitative smoking history extraction. The first stage takes sentences as inputs and outputs the lexicons using regular expressions, which are subsequently used to define high-level rules in the second stage.^9^

### Pre-trained BERT Models

Pre-trained BERT models utilize a transformer architecture and are pre-trained on a large corpus with two objectives: masked language modeling and next-sentence prediction.^12^ They are also fine-tuned for specific downstream tasks, making them highly adaptable for extracting nuanced information, such as quantitative smoking history from clinical narratives in diverse datasets. We investigated four variants of BERT developed for different clinical uses:

1. Bert-Base-Uncased^12^: The original BERT model was introduced by Devlin et al., 2019. It is pre-trained on a large corpus of English text, including BooksCorpus and Wikipedia, using uncased tokenization (ignoring capitalization). The model serves as a general-purpose baseline for various NLP tasks and can be fine-tune for specific applications.
2. BioBERT^18^: A BERT model pre-trained on large-scale biomedical corpora such as PubMed and PMC, designed for tasks like biomedical NER and relation extraction.
3. ClinicalBERT^19^: A BERT variant fine-tuned on clinical text from EHRs, focusing on clinical NLP tasks like patient mortality prediction and medical event extraction.
4. MedBERT^20^: Designed for structured healthcare data, such as medical codes and records, this model aims at predictive analytics and healthcare-specific NLP tasks.

### Experiment Pipeline

The pipeline of quantitative smoking history extraction consists of two steps: selection of a fine-tuned BERT-based model and model evaluation (**Figure 2**). SmokeBERT was developed in Step 1 and evaluated against the SOTA (rule-based) model on two tasks: NER and clinical tasks such as LCS eligibility. (Step 2).

**Figure 2.**
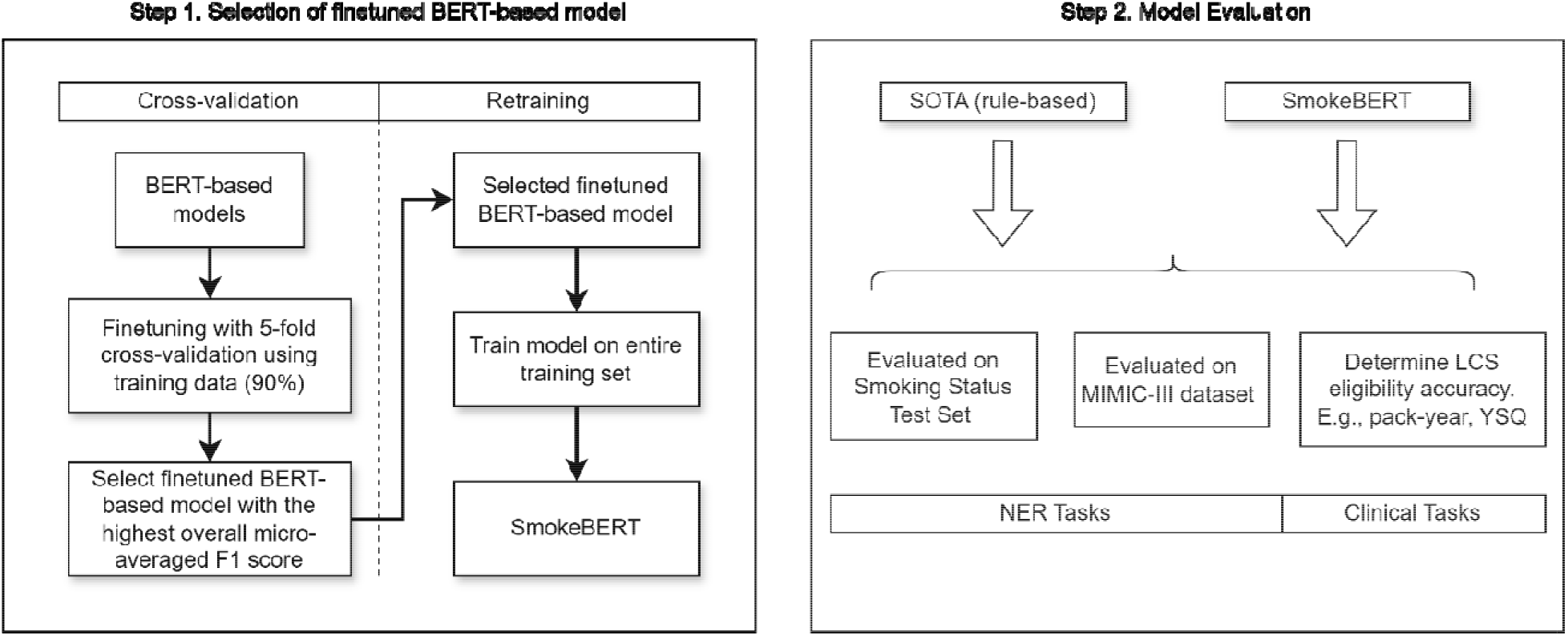
Overview of NLP pipeline. Abbreviations: BERT: Bidirectional Encoder Representations from Transformers, NER: named entity recognition, MIMIC: the Medical Information Mart for Intensive Care, LCS: lung cancer screening.

#### Step 1. Selection of Finetuned BERT-Based Model

SmokeBERT was developed after a 2-stage training, cross-validation training, and entire dataset retraining. In cross-validation training, we fine-tuned four pre-trained BERT-based models—BERT, BioBERT, ClinicalBERT, and MedBERT— using the annotated smoking history dataset. The data was split into 90% training and 10% testing, with the training portion further utilizing a 5-fold cross-validation approach. Fine-tuning was performed to optimize the model for extracting smoking-related variables. The model with the highest overall micro-averaged F1 score was selected as the final BERT variant. The overall F1 score was computed by combining results across folds and evaluated for six smoking-related variables. The BERT variant with the highest F1 score for more than 50 % of the variables (> 3 out of 6) was selected. This model was then retrained on the entire smoking history training dataset (90%) to create the final version of SmokeBERT, leveraging all available training data for improved accuracy.

#### Step 2. Model Evaluation

The evaluation pipeline involved testing SmokeBERT on both NER and LCS eligibility tasks. The hold-out 10% smoking history test set and the external MIMIC-III dataset were used for evaluating the performance of NER tasks on metrics such as micro-averaged precision, recall, and F1 score of each variable and the overall micro-averaged results. Additionally, the model’s output, consisting of structured tabular files with each sentence as a row and each smoking-related variable as a column, was evaluated for clinical applications. Specifically, tasks such as extracting pack years and years since quitting for determining LCS eligibility were assessed by calculating the proportion of correctly extracted variables against the total occurrences. For benchmarking, the same evaluation was applied to the SOTA rule-based model.

### Evaluation Metric

We evaluated smoking variable extraction at the sentence level using a two-step approach. First, performance (precision, recall, F1 score) was assessed individually for each smoking-related variable. A prediction is considered correct if the extracted value matches the ground truth annotations within the same sentence. For a single variable, if multiple conflicting values are present in a sentence—an inherently incorrect scenario since we assumed one sentence represents one patient—the extraction is considered correct only if all occurrences were captured. Second, an overall evaluation was performed by aggregating the results across all variables using a micro-averaged metric. This approach combined true positives (TPs), false positives (FPs), and false negatives (FNs) for all variables. The evaluation metrics were defined as follows:

- TP: The sentence contains a specific smoking data element, and the model correctly extracts it.
- FP: The sentence does not contain a specific smoking data element, but the model extracts some information for that specific data element.
- FN: The sentence contains a specific smoking data element, but the model extracts nothing.

### Experiment Setup

The implementation of models leverages the Hugging Face Transformers library^21^. Pre-trained BERT-based models, including MedBERT, BioBERT, and ClinicalBERT, were fine-tuned from BERT on domain-specific datasets to address our NER task. Clinical notes were preprocessed using NLTK^22^ for sentence tokenization, custom scripts were developed to label tokens for smoking-related variables. The training pipeline used the Hugging Face Trainer API for 10 epochs on the AdamW optimizer with a learning rate of 2e-5 and weight decay of 0.01. The number of epochs (10) was determined based on the performance of the F1 score from preliminary experiments, where the model began to converge after 5 epochs and stabilized around the 10th epoch. During training, early stopping was applied and set at 3 epochs if the F1 score did not improve. Evaluation metrics, including precision, recall, and F1 score, were calculated using the Seqeval package.^23^

The SmokeBERT was implemented in Python 3.5 using PyTorch (version 0.4.1). The fine-tuning and testing of the BERT-based models were conducted on one NVIDIA Quadro RTX 8000 with onboard GPU memory. The evaluation of the SOTA model was conducted on the CPU.

## Results

### SmokeBERT

**Table 2** summarizes the performance of the four fine-tuned models (BERT, BioBERT, MedBERT, ClinicalBERT) across six variables and their overall results for the 5-fold validation sets during model training. For the evaluation of this and the following table, we primarily focus on F1 scores as a primary metric to capture the models’ ability to accurately and comprehensively extract smoking-related data. Notably, recall follows a similar trend to the F1 score. ClinicalBERT achieved the best performance in F1 scores, excelling in three out of six variables and demonstrating strong recall in three out of six variables. On overall micro-averaged recall and F1 scores, ClinicalBERT reaches 0.94 and 0.93 separately, which is also the best compared to other models. Based on these results, ClinicalBERT is selected for further refinement and retraining on the complete smoking history training dataset, resulting in the SmokeBERT model.

**Table 2.** Performance comparison of finetuned models across six variables for the 5-fold validation sets in model.

| Variable | Number of Entities | Model (finetuned) | Recall | Precision | F1 |
| --- | --- | --- | --- | --- | --- |
| Packs per day | 110 | BERT | 0.93 (0.05) | 0.83 (0.03) | 0.88 (0.02) |
|  |  | BioBERT | **0.95 (0.08)** | **0.87 (0.05)** | **0.90 (0.05)** |
|  |  | MedBERT | 0.91 (0.07) | 0.77 (0.05) | 0.83 (0.05) |
|  |  | ClinicalBERT | 0.89 (0.07) | 0.79 (0.06) | 0.84 (0.06) |
| Cigs per day | 56 | BERT | **0.95 (0.10)** | 0.85 (0.11) | **0.90 (0.10)** |
|  |  | BioBERT | 0.93 (0.10) | 0.85 (0.10) | 0.89 (0.09) |
|  |  | MedBERT | 0.91 (0.28) | 0.81 (0.22) | 0.86 (0.25) |
|  |  | ClinicalBERT | 0.91 (0.19) | **0.86 (0.19)** | 0.89 (0.19) |
| Pack Years | 1244 | BERT | 0.93 (0.01) | 0.89 (0.02) | 0.91 (0.01) |
|  |  | BioBERT | **0.95 (0.01)** | **0.93 (0.02)** | **0.94 (0.02)** |
|  |  | MedBERT | 0.94 (0.01) | 0.91 (0.01) | 0.92 (0.01) |
|  |  | ClinicalBERT | **0.95 (0.02)** | **0.93 (0.03)** | **0.94 (0.02)** |
| Quit in the year | 76 | BERT | 0.75 (0.09) | 0.71 (0.11) | 0.73 (0.09) |
|  |  | BioBERT | 0.75 (0.14) | 0.68 (0.09) | 0.71 (0.08) |
|  |  | MedBERT | **0.82 (0.12)** | **0.74 (0.07)** | **0.78 (0.03)** |
|  |  | ClinicalBERT | 0.74 (0.17) | 0.70 (0.13) | 0.72 (0.14) |
| Years Smoked | 179 | BERT | 0.94 (0.01) | **0.86 (0.05)** | **0.90 (0.02)** |
|  |  | BioBERT | 0.93 (0.03) | **0.86 (0.04)** | 0.89 (0.04) |
|  |  | MedBERT | 0.93 (0.02) | 0.85 (0.07) | 0.89 (0.05) |
|  |  | ClinicalBERT | **0.95 (0.02)** | 0.85 (0.03) | **0.90 (0.02)** |
| YSQ | 408 | BERT | 0.96 (0.02) | 0.93 (0.04) | 0.94 (0.03) |
|  |  | BioBERT | 0.96 (0.03) | 0.93 (0.04) | 0.95 (0.03) |
|  |  | MedBERT | 0.95 (0.03) | 0.93 (0.05) | 0.94 (0.04) |
|  |  | ClinicalBERT | **0.97 (0.02)** | **0.95 (0.02)** | **0.96 (0.01)** |
| Overall (micro-averaged) | 2073 | BERT | 0.93 (0.01) | 0.89 (0.01) | 0.91 (0.01) |
|  |  | BioBERT | **0.94 (0.01)** | **0.91 (0.02)** | **0.93 (0.01)** |
|  |  | MedBERT | 0.93 (0.01) | 0.89 (0.02) | 0.91 (0.02) |
|  |  | ClinicalBERT | **0.94 (0.01)** | **0.91 (0.01)** | **0.93 (0.01)** |

Notes: The best-performing model for each variable and performance metric is highlighted in bold. Abbreviations: YSQ: years since quitting; BERT: Bidirectional Encoder Representations from Transformers.

### NER Task Evaluation

Figure 3. and **Table S2** in the supplement compare the performance of the SOTA rule-based model and SmokeBERT on the 10% smoking history test set. SmokeBERT significantly outperformed the SOTA model, achieving higher recall and F1 scores across all variables: recall and F1 scores consistently exceeded 0.90 and approached 1 for most variables. For precision, SmokeBERT achieves values ranging from 0.9 to 1. Recall demonstrated a similar trend to F1 scores. All metrics for the variable “Cigs per day” in the SOTA rule-based model are 0, as this model is not designed to extract this variable. Figure 4. and **Table S3** in the supplement present the performance of SmokeBERT and the SOTA rule-based model on the MIMIC-III dataset. SmokeBERT outperformed the SOTA model in recall and F1 scores across all six variables. However, for “Packs per day” and “Years Smoked”, the SOTA model achieved slightly higher scores on precision. Additionally, SmokeBERT’s recall and F1 scores for ‘Quit in the year’ were relatively low, at 0.25 and 0. , respectively, indicating room for further improvement.

**Figure 3.**
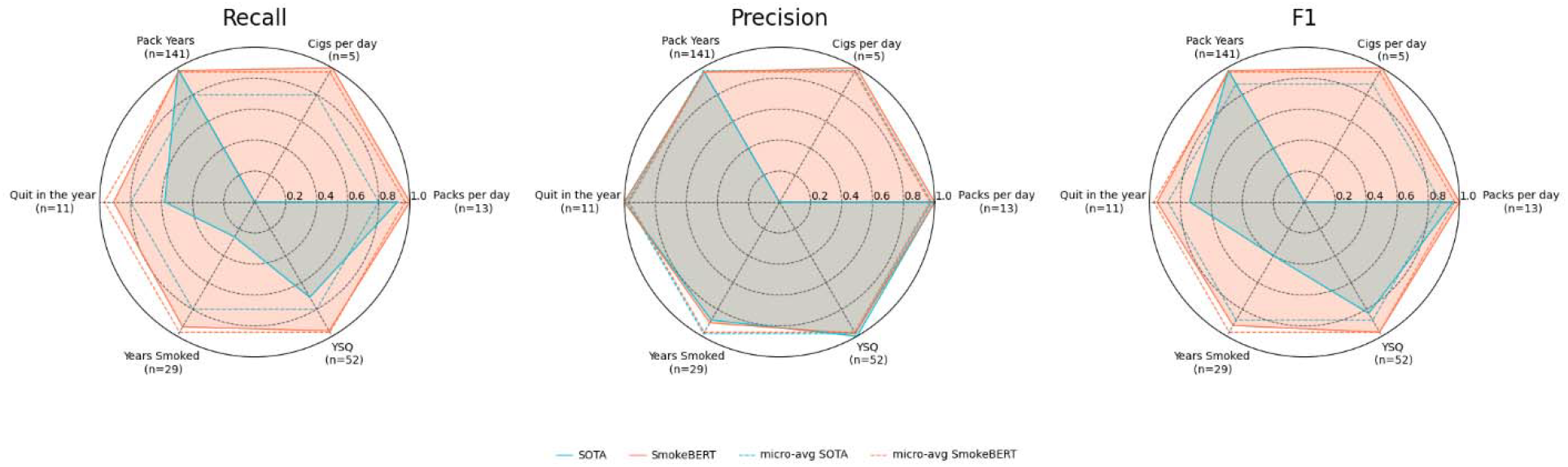
Spider charts of model performance on 10 % hold-out (smoking history) test set. Abbreviations: SOTA: state-of-the-art; YSQ: year since quitting; BERT: Bidirectional Encoder Representations from Transformers.

**Figure 4.**
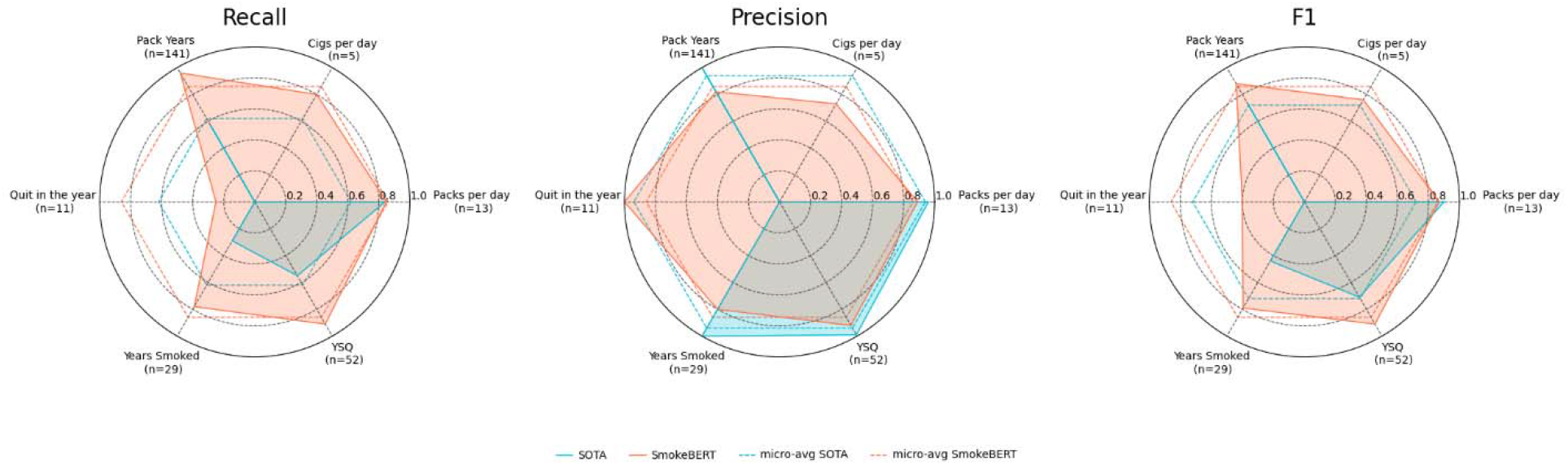
Spider charts of model performance on the MIMIC-III dataset. Abbreviations: MIMIC-III: the Medical Information Mart for Intensive Care III; SOTA: state-of-the-art; YSQ: years since quitting; BERT: Bidirectional Encoder Representations from Transformers.

### Clinical Task Evaluation

We also evaluated the performance of SOTA and SmokeBERT on the clinical tasks, such as identifying patients who meet LCS eligibility criteria based on smoking history (**Table 3**). SmokeBERT demonstrates a substantial improvement over the SOTA model in identifying patients meeting LCS eligibility criteria. For the ≥20 pack years criterion, SmokeBERT achieves a 98% success rate compared to 59.4% (60 out of 101) of SOTA, correctly identifying 98 out of 101 cases. Similarly, for the ≤15 YSQ criterion, SmokeBERT achieves 100% (87 out of 87) accuracy, outperforming SOTA’s 59.7% (52 out of 87).

**Table 3.** Model performance on clinical tasks.

| Model | % Met LCS eligibility criterion: $\geq 20$<br>pack years (n=101) | % Met LCS eligibility criterion: ≤<br>15 YSQ (n=87) |
| --- | --- | --- |
| SOTA | 59.4 (60) | 59.7 (52) |
| SmokeBERT | 97.0 (98) | 100 (87) |

Abbreviations: LCS: lung cancer screening; SOTA: state-of-the-art; YSQ: years since quitting; BERT: Bidirectional Encoder Representations from Transformers.

### Error Analysis of Clinical Task Evaluation

The SOTA model was overly sensitive to variations in the format of the unit of pack years and failed to capture many occurrences when the unit format was not included in the model’s rules, such as “PK/YR,” “PKG/YEAR,” “p yr,” and “pks/yr.” On the other hand, SmokeBERT was able to capture most variations in the unit of pack years, achieving very high performance in the clinical task evaluation. However, there were three entities, “50ppy”, “70ppy”, and “100ppyhx” that both models failed to extract, suggesting potential room for improvement in SmokeBERT’s generalizability by incorporating a wider range of sentence examples to better capture variations in the format of pack year units.

## Discussion

Large language models have been applied to classify smoking status.^24^ Yet models capable of extracting quantitative smoking information from clinical narratives remain scarce. Our SmokeBERT model demonstrated substantial improvements over the SOTA rule-based model across diverse datasets and tasks in extracting quantitative smoking history from clinical notes. During training, BioBERT outperformed other models and was refined into SmokeBERT. On test datasets, SmokeBERT consistently surpassed the SOTA model, achieving high recall and F1 scores for most variables. In clinical tasks, SmokeBERT also excelled in identifying patient eligibility for LCS.

The results highlight SmokeBERT’s potential for enhancing clinical workflows and research tasks of smoking-related data extraction. Its strong performance on the smoking history test set underscores its reliability and accuracy in extracting key smoking data elements. SmokeBERT’s superior performance in clinical tasks, such as identifying patients eligible for LCS, demonstrates its practical utility in improving clinical decision-making and ensuring accurate patient stratification, ultimately supporting efforts to address the persistently low uptake of LCS across the nation. These findings emphasize SmokeBERT’s value in advancing the extraction of smoking-related data for both research and healthcare applications.

SmokeBERT’s performance on the MIMIC-III dataset also highlights certain areas for improvement. The model struggled with variables like “Packs per day” and “Quit in the Year,” where either SOTA performed slightly better or no metrics were extracted. Additionally, SmokeBERT’s lower precision on MIMIC-III compared to the smoking history dataset suggests challenges in generalizing to datasets with varied distributions or more complex representations. These limitations highlight the need for further refinement to improve SmokeBERT’s robustness and adaptability across diverse clinical datasets.

This study has several limitations. First, patient-level information was not available. However, our dataset includes a large and diverse sample of clinical notes, which provides substantial variability in the language used to describe smoking history. We also reduced redundancy by removing duplicate sentences. Second, smoking history sentences were identified using keyword matching, which may have introduced selection bias in the training and testing datasets if relevant sentences were missed. Nonetheless, we believe the current keyword set captures most of the smoking-related documentation, as it is unlikely for smoking history to be recorded without the use of terms derived from the root “smoke,” which was included in the extraction criteria.

Future directions for enhancing SmokeBERT focus on expanding its scope, improving clinical relevance, and addressing text processing challenges. To enable global applicability, incorporating multilingual datasets, such as those in Chinese, Spanish, and French, is essential. Developing language-agnostic approaches and exploring ensemble methods will also enhance its scalability and performance in diverse settings. Testing SmokeBERT on larger and more diverse clinical datasets is critical for ensuring robustness across various healthcare contexts. Additionally, adapting models like SentenceBERT to handle clinical notes, which are often structured in paragraphs rather than sentences, will improve data extraction accuracy. This includes implementing steps to automatically segment paragraphs into meaningful sentences for patient-specific analysis. Furthermore, text augmentation techniques can be utilized to enrich training data, addressing issues such as typos due to keyboard proximity (e.g., confusing “a” and “s”) and handling synonym variability. These strategies will help refine SmokeBERT’s ability to generalize and improve its reliability for extracting smoking-related data in global and clinical contexts.

## Conclusion

In this study, we developed SmokeBERT, a fine-tuned BERT-based model for extracting quantitative smoking histories from clinical narratives. By leveraging pre-trained BERT models such as BioBERT, ClinicalBERT, and MedBERT, we addressed the limitations of existing rule-based NLP models, including their reliance on manually crafted rules and poor generalizability. Our evaluation demonstrated that SmokeBERT outperforms the SOTA rule-based model in recall and F1 scores across multiple datasets and variables. SmokeBERT also significantly improved performance in clinical tasks, such as identifying patients eligible for LCS, demonstrating its potential for enhancing clinical decision-making. Future research for SmokeBERT will investigate multilingual expansion through diverse language datasets and language-agnostic architectures to enable global deployment. Additionally, sentenceBERT adaptations for clinical note processing will enhance extraction accuracy from complex healthcare documentation. Finally, text augmentation techniques targeting typos and synonyms will improve model generalizability.

## Data Availability

The code of this study is available at: https://github.com/Elena145/SmokeBERT.

## Acknowledgments

The code of this study is available at: https://github.com/Elena145/SmokeBERT. This study was supported by the 2024–2025 Jonsson Comprehensive Cancer Center Fellowship Award at the University of California, Los Angeles, and by NIH/National Cancer Institute grants U2C CA271898 and U01 CA233370.

