## Supplemental Materials for "SmokeBERT: A BERT-based Model for Quantitative Smoking History Extraction from Clinical Narratives to Improve Lung Cancer Screening"

**Supplemental Methods**

**Table S1**. Inter-rater agreement for quantitative smoking history annotation (two raters).

**Table S2.** Model performance on 10 % hold-out (smoking history) test set.

**Table S3.** Model performance on the MIMIC-III dataset.

**Supplemental Methods**

Inter-rater agreement was assessed on 300 randomly selected sentences from the smoking history dataset by two raters in two phases. First, agreement on the presence or absence of a specific smoking variable in a sentence was evaluated. Sentences containing the smoking variable were coded as 1, while those that did not mention it were coded as 0. Agreement exceeded 0.95 for all six smoking variables (**Table S1** in the Supplement). Next, among the sentences where both raters extracted a value, a two-way mixed-effects model with single-rater, absolute agreement intraclass correlation coefficient (ICC) was used to assess the reliability of the extracted smoking history measures. The ICCs for all smoking variables were above 0.96 (**Table S1** in the Supplement), indicating excellent reliability.^1^

| **Table S1. Inter-rater agreement for quantitative smoking history annotation.** | | | |
| --- | --- | --- | --- |
| Variable | Agreement on presence/absence (n=300) | Two-way mixed effects model, single rater absolute ICC | |
|  |  | n | Value (95% CI) |
| Packs per day | 0.997 | 13 | 0.995 (0.984-0.998) |
| Cigarettes per day | 1 | 2 | NA^a^ |
| Pack years | 0.953 | 232 | 0.986 (0.982-0.989) |
| Quit in the year | 0.983 | 10 | 0.996 (0.986-0.999) |
| Years smoked | 0.957 | 11 | 0.966 (0.886-0.991) |
| YSQ | 0.973 | 50 | 1 (NA) |
| Notes: ^a^ Cigarettes per day showed perfect agreement between raters, with all values identical; however, ICC could not be computed because it is undefined for variables with no variance in ratings (i.e., when all raters assign identical values across all subjects, resulting in zero total variance). | | | |
| Abbreviations: ICC: intraclass correlation coefficient; CI: confidence interval; NA: not available; YSQ: years since quitting. | | | |

| **Table S2.** Model performance on 10 % hold-out (smoking history) test set. | | | | | |
| --- | --- | --- | --- | --- | --- |
| Variable | Number of Entities | Model | Recall | Precision | F1 |
| Packs per day | 13 | SOTA | 0.92 | 1 | 0.96 |
|  |  | SmokeBERT | 1 | 1 | 1 |
| Cigs per day | 5 | SOTA | 0 | 0 | 0 |
|  |  | SmokeBERT | 1 | 1 | 1 |
| Pack Years | 141 | SOTA | 0.99 | 0.97 | 0.98 |
|  |  | SmokeBERT | 0.98 | 0.97 | 0.98 |
| Quit in the year | 11 | SOTA | 0.58 | 1 | 0.74 |
|  |  | SmokeBERT | 0.91 | 1 | 0.95 |
| Years Smoked | 29 | SOTA | 0.26 | 0.88 | 0.40 |
|  |  | SmokeBERT | 0.93 | 0.90 | 0.92 |
| YSQ | 52 | SOTA | 0.71 | 1 | 0.83 |
|  |  | SmokeBERT | 0.96 | 0.98 | 0.97 |
| Overall (micro-averaged) | 251 | SOTA | 0.80 | 0.98 | 0.88 |
|  |  | SmokeBERT | 0.97 | 0.97 | 0.97 |
| Abbreviations: SOTA: state-of-the-art; YSQ: years since quitting; BERT: Bidirectional Encoder Representations from Transformers. | | | | | |

| **Table S3.** Model performance on the MIMIC-III dataset. | | | | | |
| --- | --- | --- | --- | --- | --- |
| Variable | Number of Entities | Model | Recall | Precision | F1 |
| Packs per day | 400 | SOTA | 0.84 | 0.96 | 0.90 |
|  |  | SmokeBERT | 0.85 | 0.89 | 0.87 |
| Cigs per day | 30 | SOTA | 0 | 0 | 0 |
|  |  | SmokeBERT | 0.80 | 0.73 | 0.76 |
| Pack Years | 115 | SOTA | 0.57 | 1 | 0.73 |
|  |  | SmokeBERT | 0.96 | 0.82 | 0.88 |
| Quit in the year | 4 | SOTA | 0 | 0 | 0 |
|  |  | SmokeBERT | 0.25 | 1 | 0.40 |
| Years Smoked | 148 | SOTA | 0.29 | 1 | 0.44 |
|  |  | SmokeBERT | 0.78 | 0.80 | 0.79 |
| YSQ | 153 | SOTA | 0.55 | 0.99 | 0.71 |
|  |  | SmokeBERT | 0.91 | 0.92 | 0.91 |
| Overall (micro-averaged) | 850 | SOTA | 0.62 | 0.94 | 0.72 |
|  |  | SmokeBERT | 0.86 | 0.86 | 0.86 |
| Abbreviations: MIMIC-III: the Medical Information Mart for Intensive Care III; SOTA: state-of-the-art; YSQ: years since quitting; BERT: Bidirectional Encoder Representations from Transformers. | | | | | |
